# Resuming In-Person Classes under COVID-19: Evaluating Assigned Seating Protocols in Limiting Contacts at Postsecondary Institutions

**DOI:** 10.1101/2021.05.07.21256844

**Authors:** Daniel Daly-Grafstein, Patricia Daly, Reka Gustafson

**Affiliations:** Department of Statistics, University of British Columbia; Vancouver Coastal Health; British Columbia Centre of Disease Control

**Keywords:** COVID-19, contacts, universities, assigned seating, simulation

## Abstract

In order to limit the spread of COVID-19, Canadian postsecondary institutions are offering the majority of classes online for the 2020-21 academic year. The goal of Canada’s public health pandemic response is to reduce severe illness and mortality from COVID-19 while minimizing social disruption. To achieve this goal, post secondary institutions need practical tools to limit COVID-19 spread and facilitate contact tracing while returning students to in-person instruction. In this paper, we explore the impact of assigned seating for students attending in-person classes in reducing potential contacts. We conduct a variety of seating simulations using student enrollment data and measure the number of potential contacts under each scenario. We find that assigning seats to students significantly reduces the expected number of contacts relative to random seating, making the return to in-person classes more feasible under these scenarios.

## 1. Introduction

COVID-19 is a respiratory infection caused by the SARS-CoV-2 virus, first identified as a cause of human illness in December 2019 (Zhu et al. 2020). It is spread primarily through respiratory droplets, which are small particles >5-10 um in diameter that are expelled when a person talks, coughs, sings or sneezes. Infection can occur when a susceptible person is within 1-2 metres of an infected person, and expelled droplets enter the mouth, nose or eyes of the susceptible contact. Less commonly, infection may occur if a susceptible person touches a surface or object recently contaminated with the respiratory droplets of an infected person, and then touches their eyes, nose or mouth (World Health Organization 2020b).

Early identification of cases of COVID-19 and follow-up and quarantine of their close contacts is the most effective intervention to prevent spread of the virus. Because the incubation period (time from exposure to onset of illness) for COVID-19 is typically 5-7 days and up to 14 days, public health officials can identify and quarantine contacts before those who may have been infected are themselves infectious to others (World Health Organization 2020a). Strategies to limit the number of potential contacts in a variety of settings, including workplaces and schools, are typically included in COVID-19 safety plans. While some COVID-19 safety protocols for postsecondary institutions have included a recommendation for assigned seating in classroom settings (e.g. WorkSafeBC 2020), no analysis has been done to demonstrate how this might reduce the number of potential contacts of COVID-19 cases.

Most postsecondary institutions in Canada have decided to administer classes entirely online for the 2020 Fall term, and many have announced plans to do the same for the Winter 2021 terms. The goals of Canada’s pandemic response plan are to both reduce severe illness and mortality and to minimize social disruption (Public Health Agency of Canada, 2018). Hence the return to essential societal functions such as in-person education, is an explicit goal of Canada’s pandemic plan. In this paper we seek to show that implementing assigned seating in postsecondary school classes can reduce contacts to a level where returning to live classes is feasible. We do this by using student enrollment data and simulating class seatings to evaluate student contacts under a variety of simulation scenarios. We begin by describing the data and classroom structure used for these simulations. We next describe each of the seating scenarios we will be simulating. Finally, we compare the number of resulting contacts under each of these simulations.

## 2. Data

For this project we use anonymized student enrollment data from Term 1 (September - December) of the 2019-20 school year at the University of British Columbia (UBC). These data include the course sections each student attended along with information about the room size, number of meeting times per week, and number of seats available in each section. For this project we restrict our analysis to first-year students attending the UBC-Vancouver campus who enrolled in 4-6 courses in Term 1 and are part of a “Bachelor’s” program (BSc, BA, BCOMM, etc.). Notably, there are many class sections which have greater than 1 room associated with them in the data where it is not possible to know which room a given student attended. Therefore for our simulations we only consider students that enrolled in sections without any duplicates in the data. This leaves us with a total sample of 1566 first year students.

In order to simulate students sitting in a class we need to have a structure for how the seats are situated. For each section in our data we have the number of total seats available in the room, but we do not have information about the actual layout of seats. Therefore we decide to take a worst-case scenario approach and assume each class has all its seats together without any aisles. We also assume the number of rows and columns in each class are close to equal. If *r*_*i*_, *c*_*i*_, and *s*_*i*_ are the number of rows, columns, and seats in section *i*, we assume:

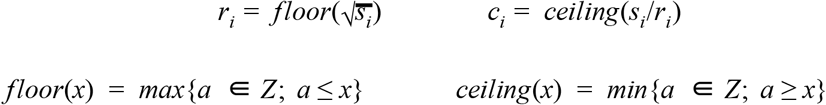

If the number of seats is not a multiple of the number of rows, the remaining seats are removed from the last row. Figure 1 shows an example seating structure for a class with 28 seats.

**Figure 1.**
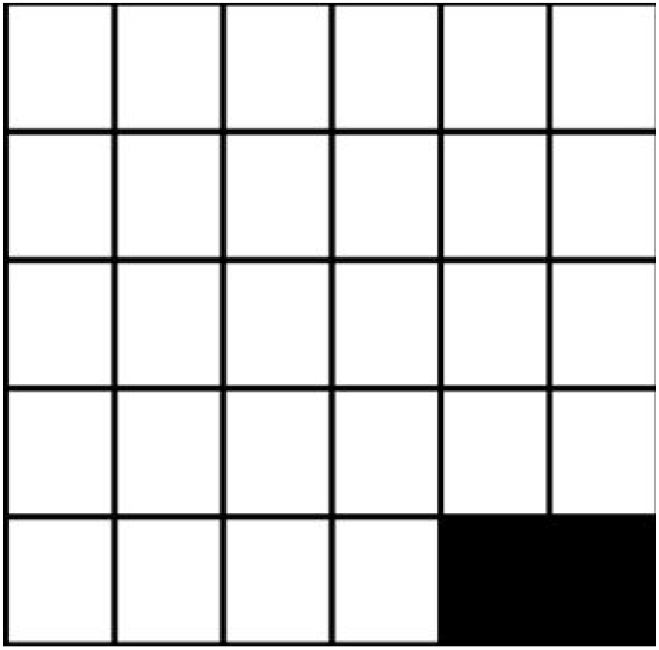
Seating structure for a class with 28 seats. This class has 5 rows and 6 columns, with the extra 2 remaining seats removed in the last row.

We define a contact for a given student as any student seated directly in front, behind, or diagonally to the given student. The maximum number of contacts for any student in a class is 8 (Figure 2).

**Figure 2.**
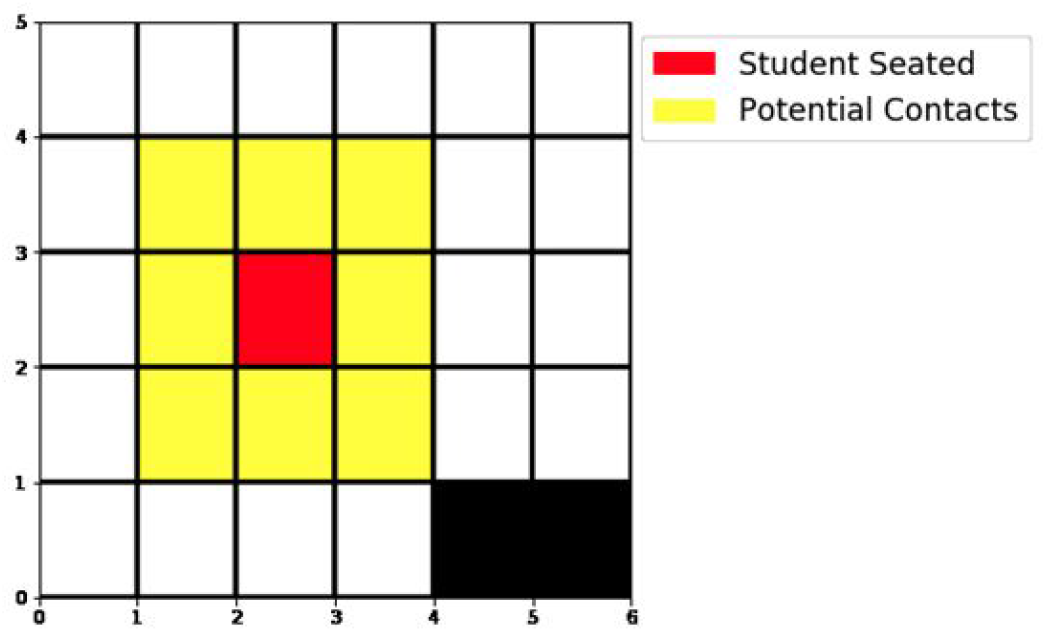
The student of interest is coloured red. The potential contacts for that student are any students seated in the seats coloured yellow.

## 3. Simulating Classes

Having defined the classroom structure for each of our sections, we describe the different scenarios under which we will conduct simulations. Simulations include all classes taken by the sample of 1566 first-year students over a two-week period (Lauer et al. 2020). We simulate all in-person meetings these students have - including lectures, tutorials, labs, seminars, and discussion classes. In Section 4 we present the results of these simulations scenarios.

### 3.1 Random Seating

We begin with a null case where students pick seats in each class completely randomly. We do this to give us a baseline to compare our other scenarios. For example, at UBC in the fall term of the 2019-20 academic year, MATH100 Section 106 meets on Tuesdays and Thursdays. Thus for the two week simulation window we simulate seatings for this section 4 times (Figure 3).

**Figure 3.**
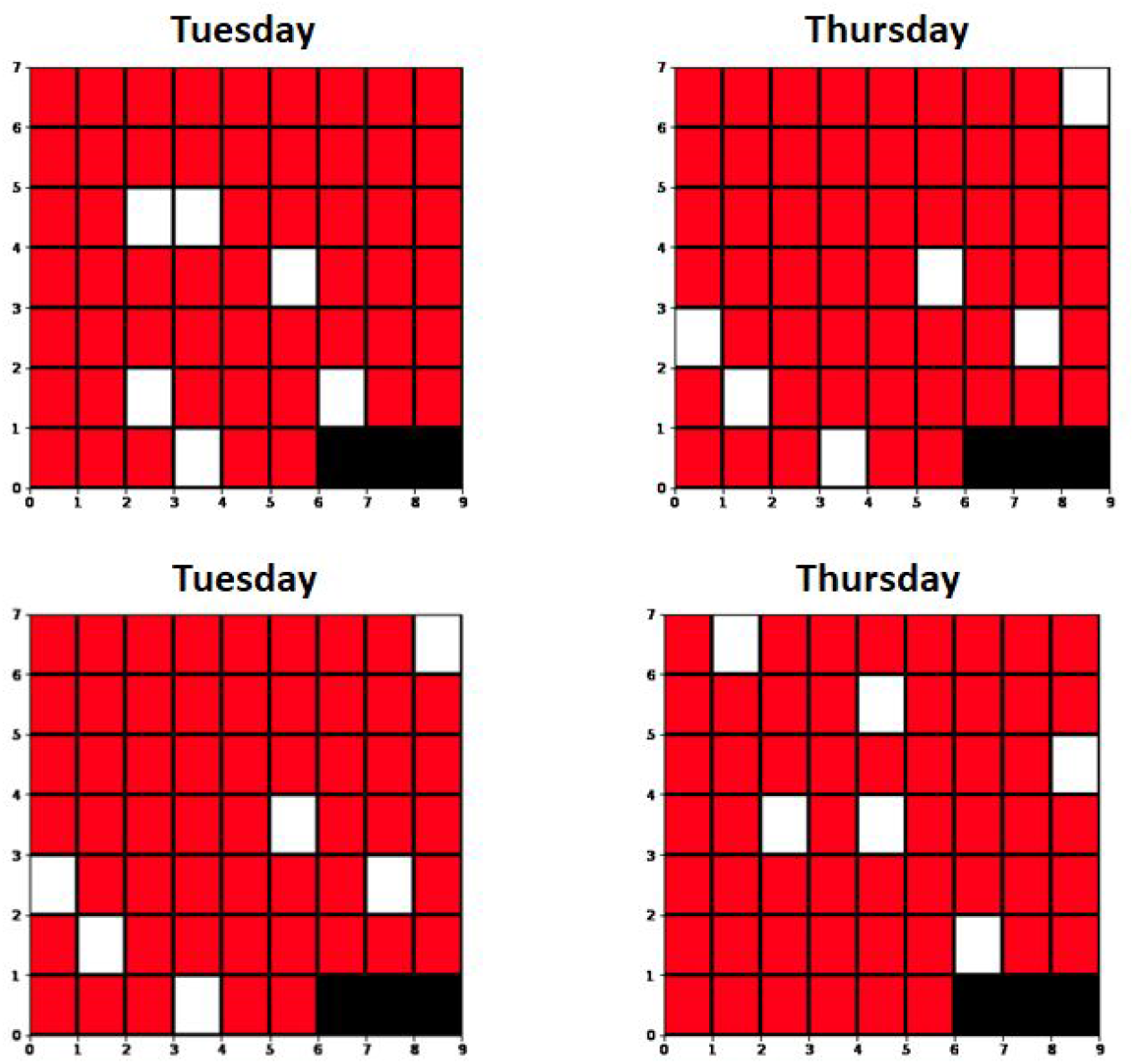
Randomly selected seats for two weeks of MATH100 Section 106. This section has 54 enrolled students and 60 seats. Red squares denote occupied seats, white squares denote empty seats, and black squares denote removed seats that cannot be occupied.

### 3.2 Realistic Seating

In our second scenario we try to more realistically simulate how students choose their seats. Students do not pick seats randomly, they prefer to sit in certain areas of the class. They also do not always attend class alone, often sitting with groups of friends. Students also generally prefer to leave empty seats between themselves and others if given the opportunity. Based on these observations we introduce the following criteria in this simulation:

For each class:

- Students are randomly assigned a quadrant to sit in for each class. These are assigned with equal probability and can be different for the same student across multiple classes. Students remain in these quadrants for the entire two-week period.
- Students are randomly assigned to sit alone or in pairs for each class with equal probability. Pairs of students always sit next to each other for a given class.
- Individuals and pairs of students will first fill seats where they can avoid sitting beside other students in the same row.

Figure 4 shows an example of how students pick seats under these conditions.

**Figure 4.**
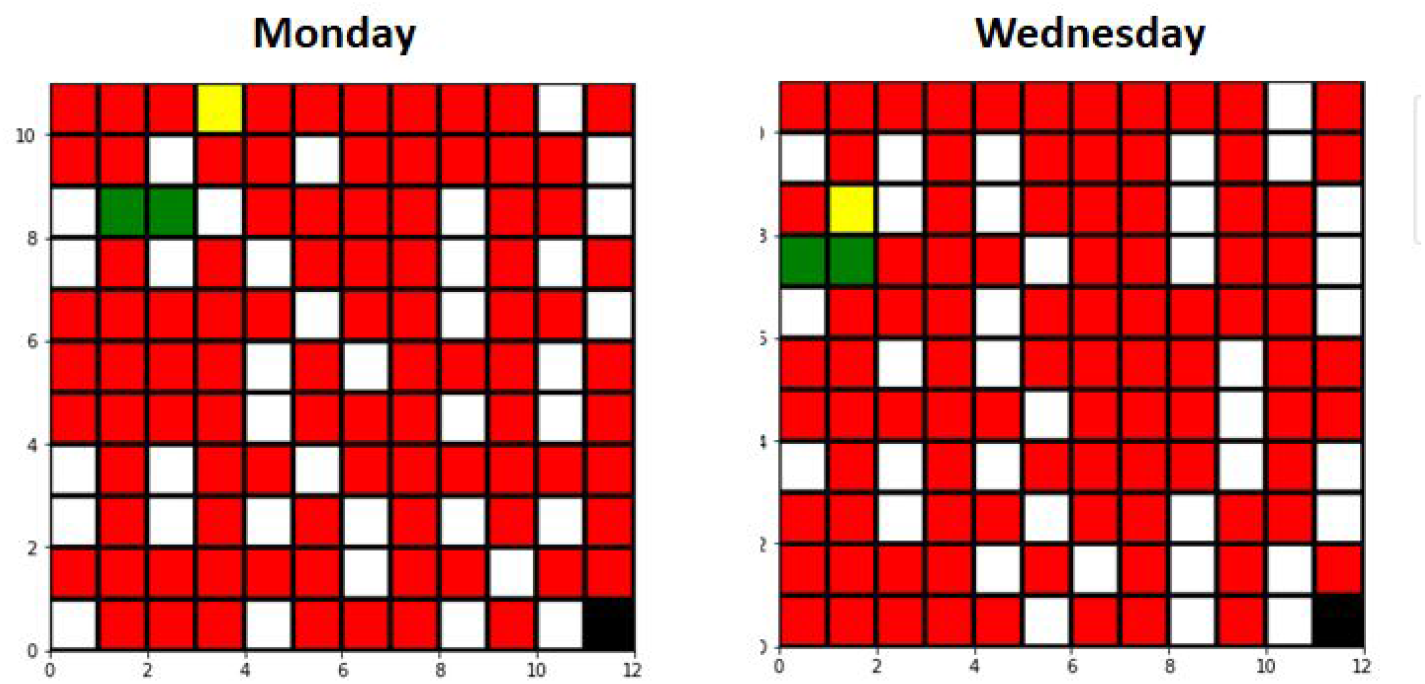
Realistically selected seats for one week of PHIL101 Section 001. This section has 94 enrolled students and 131 seats. Red squares denote occupied seats, white squares denote empty seats, black squares denote removed seats that cannot be occupied. The yellow square is an example of a student assigned to the front-left quadrant and sitting alone. The green squares are an example of a pair of students assigned to the front-left quadrant.

### 3.3 Assigned Seating

Here we introduce an intervention into our seating simulations in an attempt to reduce student contacts. Instead of students choosing seats completely randomly or randomly under some conditions, we assign each student a seat that they stay in for the entire semester. Assigning seats involves two steps - choosing which seats students will sit in, and assigning students to those chosen seats. Each of these steps can be done in two ways - randomly, or to minimize contacts between students. We will perform a total of three assigned seating simulations based on these steps: randomly choosing and assigning seats, choosing seats to minimize contacts and randomly assigning seats, and finally choosing and assigning seats to minimize contacts.

#### 3.3.1 Randomly choosing and assigning seats

Randomly choosing and assigning seats is equivalent to the scenario where students randomly picked seats for each class, except in this case the simulation is only done once for a two-week period since contacts will remain the same for every class. For example, if a class meets on Tuesdays and Thursdays, simulations in Section 3.1 will be run 4 times, while simulations here will be run once.

#### 3.3.2 Choosing seats to minimize contacts

We now attempt to choose seats in each class to minimize the number of contacts students will have. The method to choose seats can be seen in Figure 5 and is described in Algorithm 1 below:

**Figure 5.**
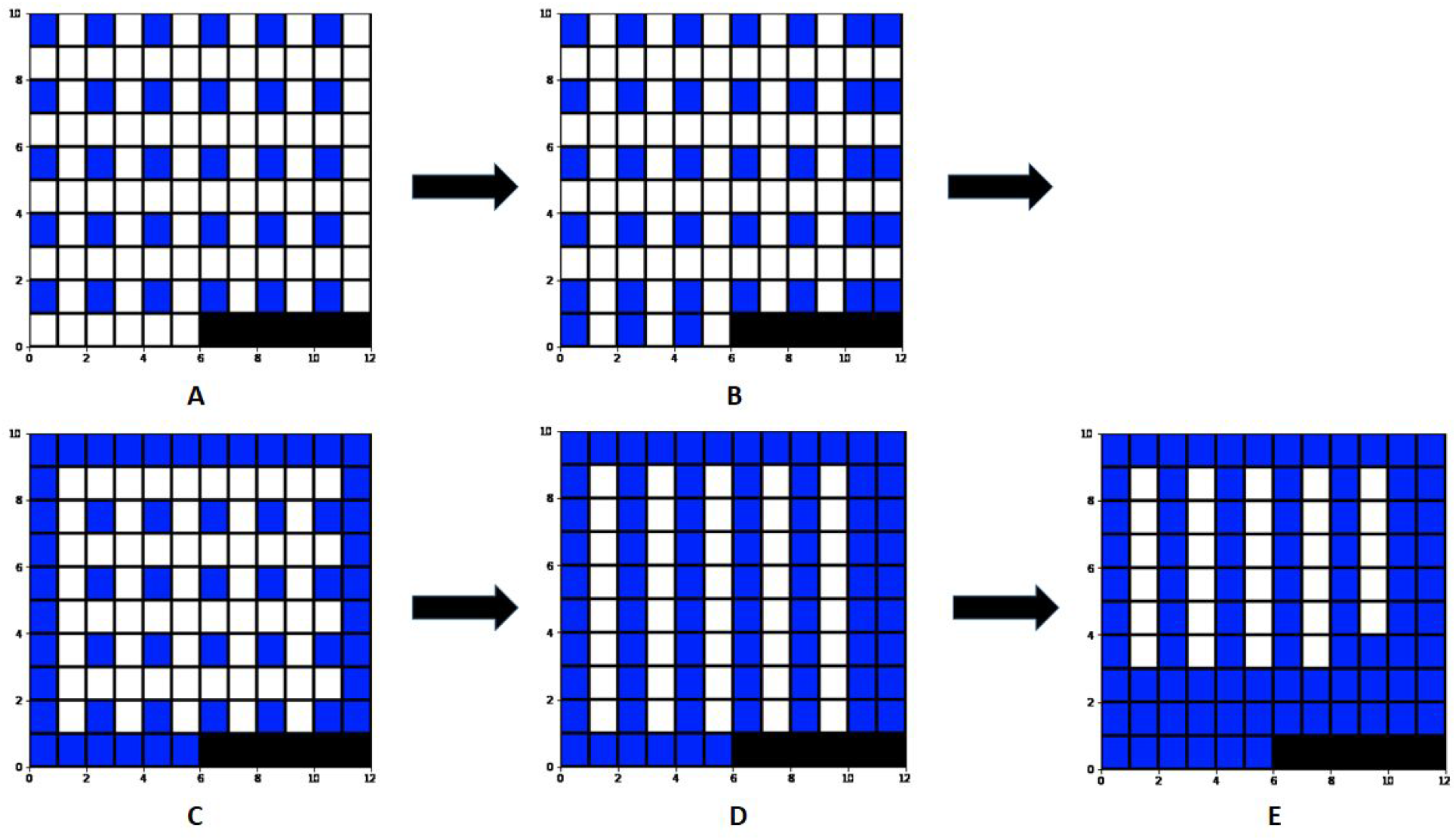
Visualizing Algorithm 1 used to choose seats in Section 3.3.2. Figures A-E correspond to steps 1-5. MATH 255 Section 102 is shown with 85 enrolled students and 114 seats.

1. Fill seats in every other row and column such that there are zero potential contacts (Figure 5A).
2. If there are an even number of rows and/or columns, fill the corresponding row/column in the same way (Figure 5B).
3. Fill all seats in the first and last columns and rows (Figure 5C).
4. Fill all remaining seats in every other column (Figure 5D).
5. Fill all remaining seats beginning in the back of the class (Figure 5E).

If at any point during the above algorithm the number of seats chosen equals the number of students in the section the process is stopped. Examples of seat distributions for sections with different proportions of students are shown in Figure 6.

**Figure 6.**
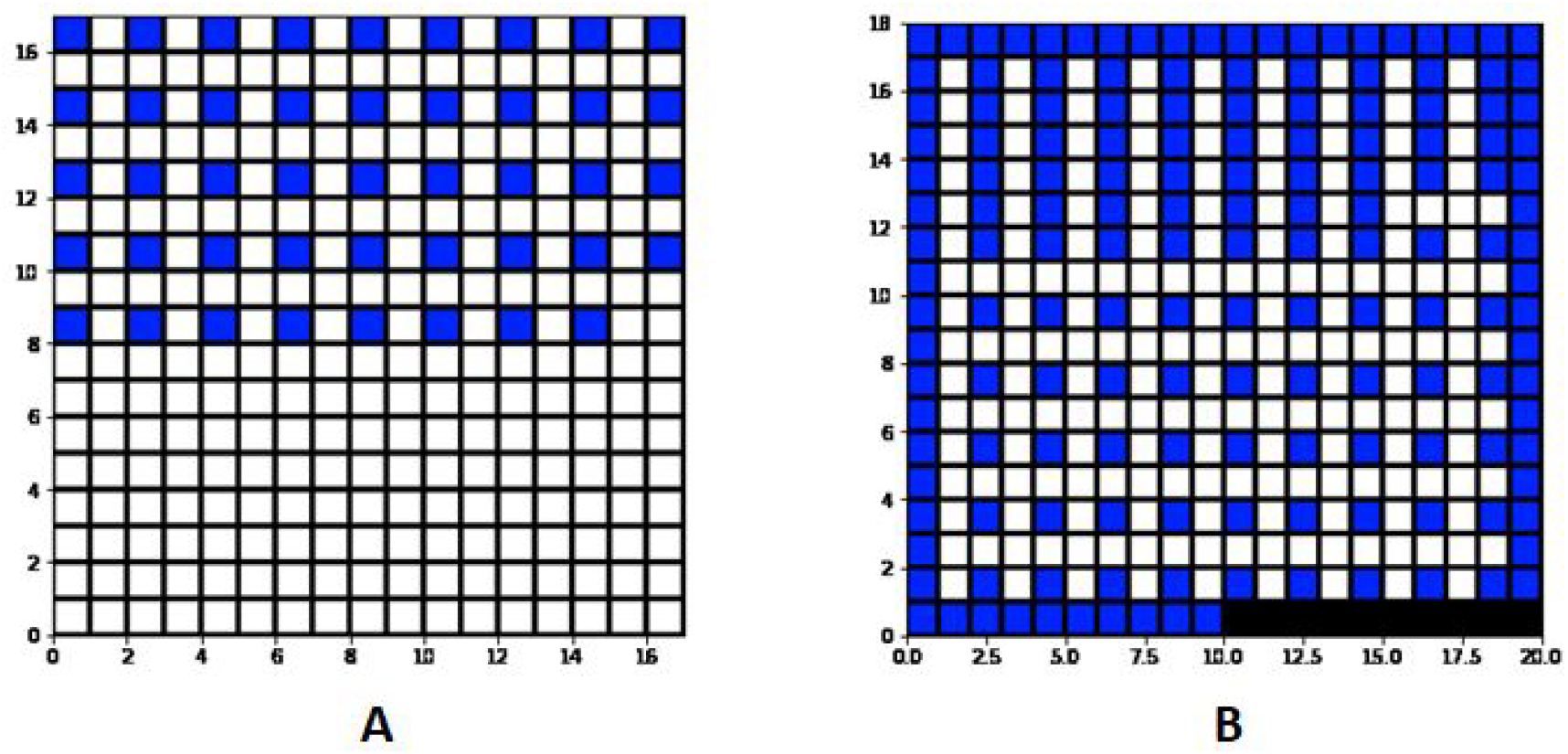
Figure A corresponds to MUSC105 Section 101 with 44 enrolled students and 289 seats. Figure B corresponds to ECON114 Section 101 with 159 enrolled students and 350 seats.

Once seats have been chosen students are randomly assigned to the chosen seats. Students remain in these seats for the entire term.

#### 3.3.3 Assigning seats to minimize contacts

Here we repeat the algorithm for choosing seats in 3.3.2 except instead of assigning seats randomly we assign seats to minimize contacts. Given we’ve chosen seats for a given class, we assign seats iteratively from the seat with the least potential contacts to the most potential contacts described in Algorithm 2 below:

1. For a given seat, get students in contact with that seat.
2. From the students that haven’t been seated, pick the student with the most total sections shared with the students in contact.
3. If tied, pick the student with the least total sections shared with the entire class.
4. If tied, pick from the tied group at random.

We note that Step 3 chooses the student with the least sections shared with the class because seats are assigned from least to most potential contacts.

## 4. Simulation Results

For each of the scenarios described in Section 3 we simulate two-weeks of classes and count each student’s unique contacts (Table 1). We find that when students choose seats completely randomly they average 129.74 unique contacts, when students pick seats more realistically as described in Section 3.2 they average 98.86 unique contacts, and when students are assigned seats they average less than 40 unique contacts over a two-week period.

**Table 1.** Simulated mean, max, and total contacts for a sample of 1566 first-year UBC students under various simulation criteria. ‘Min choose’ refers to choosing seats according to Algorithm 1, ‘min assign’ refers to assigning seats according to Algorithm 2. The last two rows refer to simulations described in Section 4, removing all classes greater than 300 students or 200 students, respectively.

| Seating Method | Mean Contacts | Max Contacts | Total Contacts |
| --- | --- | --- | --- |
| Random Seating | 129.74 | 222 | 203,171 |
| Realistic Seating | 98.86 | 192 | 154,820 |
| Assigned seating – randomly choose, randomly assign | 39.75 | 73 | 62,244 |
| Assigned Seating – min choose, randomly assign | 33.05 | 72 | 51,764 |
| Assigned Seating – min choose, min assign | 30.52 | 70 | 47,789 |
| Assigned Seating – min choose, randomly assign, exclude >300 | 26.10 | 61 | 40,888 |
| Assigned Seating – min choose, randomly assign, exclude >200 | 19.85 | 53 | 31,096 |

We can add modifications to the above simulations to try and further reduce student contacts. Here we try removing classes over a certain size, modifying the simulations done in Section 3.3.2. We repeat the simulation twice - removing all classes with greater than 300 students enrolled, and greater than 200 students enrolled, respectively. This simulation represents a mixed approach where students attend some live classes but larger classes remain online. We find removing these larger sections significantly reduces the average number of contacts (Table 1). There is a trade-off in this mixed approach - removing many sections can significantly reduce contacts, but at some point the majority of students will have most of their classes online.

Table 2 summarizes the number of live sections students have under these different approaches. We can see that when removing classes with greater than 300 students 92% of the students in our sample still have 3 or more live courses.

**Table 2.** Number of students with 0-6 live courses with all sections attended, sections with greater than 300 students removed, and sections with greater than 200 students removed. Note our sample only includes students who took 4-6 courses in the term.

| Number of Live Sections | Zero | One | Two | Three | Four | Five | Six |
| --- | --- | --- | --- | --- | --- | --- | --- |
| No Classes Removed | 0 | 0 | 0 | 0 | 708 | 602 | 256 |
| >300 person classes removed | 2 | 20 | 106 | 360 | 715 | 318 | 45 |
| >200 person classes removed | 73 | 196 | 459 | 465 | 265 | 85 | 23 |

## 5. Conclusion

This work shows that assigning seats for postsecondary school classes has the potential to significantly reduce student contacts of infectious COVID-19 cases while still allowing gatherings far larger than the 50 people currently permitted in British Columbia. We showed through simulation studies that assigning seats, whether randomly or to minimize contacts, can reduce contacts to less than 40 per student, compared to 100 or more contacts per student when seats are chosen randomly. Additionally, many of the assumptions made for these simulations were conservative to ensure potential contacts were not undercounted. For example, students were assumed not to miss any lectures, seats were assumed to be as close together as possible, and labs, discussions, and tutorials were treated the same as lectures in terms of how students were seated. It is likely that relaxing these assumptions to allow some students to miss classes, and acquiring more detailed information about seating plans would further reduce the estimated number of contacts. An additional benefit of assigned seating is the ability to identify and contact students who were sitting next to an infectious case of COVID-19 for contact tracing and management. This additional benefit is not quantified.

As post-secondary institutions have reopened to in-person learning, clusters of COVID-19 have been reported, often associated with student gatherings and congregate living settings (Wilson et al. 2020). Implementing assigned seating to support resumption of lectures and in-class learning will bring more students to campus, potentially increasing risk of COVID-19 transmission in other settings. It is important that post-secondary institutions work with local public health officials to fully implement robust, comprehensive COVID-19 safety plans that consider risks of COVID-19 transmission in all settings.

## Data Availability

All data in this article are based on simulations.

## 6. Acknowledgements

This research was funded by the BC Centre for Disease Control.

